# Chinese medicine (Q-14) in the Treatment of Patients with Coronavirus Disease 2019 (COVID-19): A Single-center, Open label, Randomised Controlled Trial

**DOI:** 10.1101/2021.01.25.21249417

**Authors:** Jia Liu, Wei Yang, Yue Liu, Cheng Lv, Lianguo Ruan, Chen Zhao, Ruili Huo, Xin Shen, Qing Miao, Wenliang Lv, Hao Li, Huaxin Shi, Lijie Hu, Zhixu Yang, Li Zhang, Bing Wang, Guoju Dong, Yongyue Xian, Bin Li, Zhenqi Zhou, Chunyan Xu, Yingying Chen, Yongjun Bian, Jing Guo, Jinliang Yang, Jian Wang, Wensheng Qi, Suping Chen, Yang Chen, Bei Yan, Wei Wang, Jing Li, Xiaolei Xie, Ming Xu, Jianxin Jiang, Gang Wang, Xiaodong Cong, Haoning Zhu, Jiaheng Shi, Luxing Leng, Dongxu Li, Lanping Guo, Luqi Huang

**Affiliations:** China Academy of Chinese Medical Sciences, Beijing, China; China Academy of Chinese Medical Sciences Institute of Basic research in Clinical Medicine, Beijing, China; China Academy of Chinese Medical Sciences Xiyuan Hospital, Beijing, China; Wuhan Jinyintan Hospital, Wuhan, Hubei, China; China Academy of Chinese Medical Sciences Institute of Chinese Materia Medica, Beijing, China; China Academy of Chinese Medical Sciences Guanganmen Hospital, Beijing, China; China Academy of Chinese Medical Sciences Chinese Materia Medica Resource Center, Beijing, China; China Center for Evidence-based Medicine of Traditional Chinese Medicine, China Academy of Chinese Medical Sciences, Beijing, China

**Keywords:** COVID-19, Chinese medicine, efficacy, safety, Randomised Controlled Trial, Q-14

## Abstract

**OBJECTIVE:** To evaluate the efficacy and safety of Chinese medicine (Q-14) plus standard care compared with standard care alone in adult with coronavirus disease 2019 (COVID-19).

**Study DESIGN:** Single-center, open label, randomised controlled trial.

**SETTING:** Wuhan Jinyintan Hospital, Wuhan, China, February 27 to March 27, 2020.

**PARTICIPANTS:** 204 patients with laboratory confirmed COVID-19 were randomised in to treatment group and control group, which was 102 patients each group.

**INTERVENTIONS:** In treatment group, Q-14 was administrated at 10g (granules), twice daily for 14 days and plus standard care. In control group, patients were given standard care alone for 14 days.

**MAIN OUTCOME MEASURE:** The primary outcome was conversion time of SARS-CoV-2 viral assay. Adverse events were analyzed in the safety population.

**RESULTS:** Among 204 patients, 195 were analyzed according to the intention to treat principle. There were 149 patients (71 vs. 78 in treatment group and control group respectively) turning to negative via SARS-CoV-2 viral assay. No statistically significance showed in conversion time between treatment group and control group (FAS: Median (IQR): 10.00 (9.00-11.00) vs. 10.00 (9.00-11.00); Mean rank: 67.92 vs. 81.44; P=0.051.). Time to recovery of fever was shorter in treatment group as compared in control group. The disappearance rate of symptom in cough, fatigue, chest discomfort was significantly higher in treatment group. In chest computed tomography (Chest CT) examinations, overall evaluation of chest CT examination after treatment compared with baseline showed more patients improved in treatment group .There were no significant differences in the other outcomes.

**CONCLUSION:** Administration of Q-14 on standard care for COVID-19 was useful for improvement of symptoms (such as fever, cough, fatigue and chest discomfort), while did not result in a significantly higher probability of negative conversion of SARS-CoV-2 viral assay. No serious adverse events were reported.

**TRIAL REGISTRATION:** ChiCTR2000030288

## Introduction

CoronaVirus Disease 2019 (COVID-19) pandemic caused by SARS-CoV-2 has resulted in considerable morbidity and mortality in more than 200 countries, which has become a major public health crisis on a global scale (Wang et al., 2020). By August 24, 2020, the number of confirmed cases of COVID-19 has reached more than 23 million, the total reported deaths now exceeds 800,000 globally, with rapid daily increases in some countries (WHO, 2020). Information regarding the epidemiology and clinical features of COVID-19 (Wu et al., 2020; Chen et al., 2020) provides good support for its clinical prevention and treatment, but many of the drugs previously hailed as star drugs, such as remdesivir, favipiravir, lopinavir-ritonavir and chloroquine or hydroxychloroquine have not shown ideal results in clinical trials, even some of them has been found to have obvious toxic side effects (Wang et al., 2020; Tang et al. 2020). Scientists around the world are actively seeking potential effective drugs or developing vaccine to treat COVID-19.

China quickly halted the spread of COVID-19 with strong containment measures (Li et al., 2020). The strategy of integrated traditional Chinese medicine (TCM) with western medicine plays an important role in the prevention and control of COVID-19 in China, more than 90% confirmed cases have received TCM therapy (SCIO-China, 2020). At the beginning of COVID-19, China National Medical Team of Traditional Chinese Medicine of China Academy of Chinese Medical Sciences (CNMTCM-CACMS Team) went to Wuhan to carry out integrated TCM with Western medicine treatment for COVID-19 in Wuhan Jinyintan Hospital, which is one of the hospitals receiving most of COVID-19 patients. In the absence of specific drugs for an emerging infectious disease, based on the theory of TCM with thousands of years of experience in the prevention and treatment of infectious disease, in particular, accumulated clinical experience of TCM in the SARS in 2003, Chinese medicine (Q-14) was developed to clinical application for treatment of COVID-19 in Wuhan, China. Q-14, also known as *Huashi Baidu granule*, is a compound granules composed of 14

Chinese herbs (see supplementary data). The National Health Commission and the National Administration of TCM of the People’s Republic of China developed “National clinical practice guideline for COVID-19 in China” (NHC-NATCM-China guidelines) (NHC-China, 2020), Q-14 is one of the main Chinese herbal preparations for the treatment of COVID-19 in NHC-NATCM-China guidelines, which was widely used in clinical practice during the outbreak of COVID-19 in Wuhan, China. One of the important ways to improve the international recognition of Chinese herbal medicine is to evaluate the clinical efficacy and safety of Chinese herbal medicine with evidence-based medicine (Tang, 2006).

At the critical moment of the COVID-19 outbreak in Wuhan of China, we conducted a single-center, open label, randomised controlled trial to assess the efficacy and safety of Q-14 in hospitalized adults with laboratory-confirmed COVID-19.

## METHODS

### Trial Oversight

In this open-label randomised trial, we recruited patients with COVID-19 from Wuhan Jinyintan Hospital, Wuhan, China. The protocol was designed in compliance with the Declaration of Helsinki, Good Clinical Practice guidelines, and local regulatory requirements. The study protocol had been approved by the Ethics Review Committee of China Academy of Chinese Medical Sciences. The protocol has been registered in China Clinical Trial Registry website (www.chictr.org/cn/, ChiCTR2000030288). All patients had given written informed consent.

China Academy of Chinese Medical Sciences research group provided the Chinese Medicine composition and Guangdong Yifang Pharmaceutical Co.,ltdproduced the compound granules. The results of this trial are reported in accordance with CONSORT (Consolidated Standards of Reporting Trials) guidelines.

### Trial design, randomization, and procedures

This study was a single-center, randomised, parallel, open label trial of Q-14 in patients with laboratory confirmed COVID-19. Participants were enrolled by investigators working in Wuhan Jinyintan Hospital. No placebo was used, and compound granules were not masked. SAS 9.4 software was used to generate random sequences in a 1:1 ratioby an independent statistician from China Center for Evidence-Based Traditional Chinese Medicine who was not aware of the trial protocol. Two research nurses, used a mobile software program “CLINICALCRS” to apply for randomization number and assigned participants to interventions. All patients accepted standard care in accordance with the NHC-NATCM-China guidelines (version 6.0, published on February 18, 2020). Patients in treatment group were given Q-14 the day after randomization, with a dose of 10g (granules), twice daily for 14 days continuously. The day of randomization was defined as day 0 and the following days were defined as day 1 to day 14. Investigators, patients and statisticians were no masked to group assignment. The routine care and laboratory staff were unaware of the treatment. Research data was recorded using Medroad Cloud Electronic Data Capture System (Jiangsu Famous Medicine Science and Technology Ltd.) Double data entry was carried out by two different recorder. Study quality control was executed in three levels, including first level by clinical research associate, second level by independent GCP (Good Clinical Practice) staffs who did not participant in the trial, third level by clinical research management department of China Academy of Chinese Medical Sciences.

### Patients

We recruited 204 patients with laboratory confirmed COVID-19 in Wuhan Jinyintan Hospital during February 27 to March 27, 2020.

The inclusion criteria included:1) Comply with the diagnostic criteria for general type COVID-19 in the NHC-NATCM-China guidelines (version 6.0); 2) 18≤Aged≤75 years; 3) Agree to participate in the trial, and the patient, the legal guardian or the person in charge of the medical institution signed the informed consent through paper signature.

The exclusion criteria included: 1) Critical patients; 2) Patients who cannot guarantee compliance of using Q-14 during the treatment period, or patients who are difficult to take medicine by oral or nasal route; 3) Patients with severe primary respiratory disease or other pathogenic microbial pneumonia that needs to be identified with COVID-19; 4) Pregnant or parturient women including patient who have a pregnancy plan or a positive urine pregnancy test; 5) Patients with other systemic malignant diseases such as malignant tumors, mental illnesses, etc., which the researchers consider unsuitable for participation in the trial; 6) Patients who have been allergic or intolerant to taking Chinese medicine herb.

The definition of critical patients was in accordance with the disease severity of COVID-19 in “National clinical practice guideline for COVID-19 in China (version 6.0).

### Outcome and Assessment

The primary outcome in this trial was conversion time of SARS-CoV-2 viral assay. The secondary outcomes were: 1) The change of 7-point scale; 2) The rate of critical aggravation; 3) Blood routine test outcomes; 4) Blood biochemical test outcomes; 5) Fever recovery time; 6) Symptom improvement; 7) Evaluation of chest CT examination after treatment compared with baseline. Adverse events were reviewed daily to assure the safety of patients.

SARS-CoV-2 viral assay for specimens from upper respiratory tract were tested on day 0, and day 7 to 15. If the test resultwas negative, SARS-CoV-2 viral assay would be conducted at least 24 hours later to observe whether there were two consecutive reports of negative result. When there were two consecutive negative results, the patient was defined to a conversion-of-negative ending and the day of the first negative report was determined as the conversion day. The time from day 0 to conversion day was defined as the conversion time.

The 7-point scale was a seven-categoryordinal scaleto test the condition and prognosis of severity during treatment. It is consisted of the following categories: 1, not hospitalized with resumption of normal activities; 2, not hospitalized,but unable to resume normal activities; 3, hospitalized, not requiring supplemental oxygen; 4,hospitalized, requiring supplemental oxygen; 5,hospitalized, requiring nasal high-flow oxygentherapy, noninvasive mechanical ventilation, orboth; 6, hospitalized, requiring ECMO (extracorporeal membrane oxygenation), invasivemechanical ventilation, or both; and 7, death.The 7-point scale was recorded during days 1 to day 14. The change of 7-point scale was the score difference after treatment when comparing with baseline.

The classification of COVID-19 was record on day 0, 1 to 14. The classification included four categories, mild, moderate, severe and critical with the sequence of severity increasing. If the classification changed to critical during days 1 to 14, the patient would be defined as critical aggravation. The rate of critical aggravation patient would be count after treatment.

Blood routine test and blood biochemical test outcomes included complete blood cell count with differential, blood chemistry, Creative protein, hypersensitive C-reactive protein, amyloid protein, myohemoglobin, hypersensitive troponin, erythrocyte sedimentation rate, ferroprotein, D-Dimer, interleukin 6, were tested on day 0, 7, 14.

Vital signs were recorded on day 0, 1 to 14. If body temperature had a fever, which was defined as over 37.3°C, we recorded the day when it began to keep stable below 37.3°C to determining fever recovery time.

The symptom including cough, fatigue, headache, chest discomfort and sore throat was recorded onday 0 and 14.The disappearance of symptom will be recorded after treatment. The symptom improvement was evaluated by the disappearance rate of the five symptoms.

Chest computed tomography examinations were tested on day 0, 14. Two radiologists with at least 20 years working experience reviewed the CT reports and images, who was unaware of the randomization. The judgment of chest CT including: 1) Score of ground glass area; 2) Score of consolidation area; 3) Density change of ground glass imaging manifestation; 4) Density change of consolidation imaging manifestation; 5) Overall evaluation of chest CT examination after treatment compared with baseline. To count the score of lesion area, each lung was divided into 10 regions according to anatomy, which were 20 regions for both sides. If the lesion area was over 50% (contained) in one region, it would be judged as 2 point, and if below 50%, it would be judged as 1 point. The range of sores for area was 0 to 40 points. Density change was recorded after treatment as decreased, no-change or increased. Overall evaluation of chest CT examination was the case and proportion of improved, no-change, and aggravated patients in each group.

In this trial, we recorded the timing, duration, severity, management and consequence of adverse events, and determined the association with the usage of study medications. (Annex 1)

### Statistical analysis

The sample size was calculated based on the alternative hypothesis that the hazard ratio of SARS-CoV-2 viral assay negative conversion rate between treatment group and control group during the 14-day trial period was *θ* ≠ 1. It was assumed that the SARS-CoV-2 viral assay negative conversion time followed an exponential distribution. We assumed that the SARS-CoV-2 viral assay negative conversion rate was 0.6 in all patients within 14 days, and the hazard ratio *θ* =2 based on the experience of treatment at the time. The sample size distribution between treatment group and control group was 1:1, the one-side superiority type I error was 0.05, and the type II error was 0.2, that is, the power was 80%. We estimated a total sample size of 204 at the drop-out rate of 15%.

For primary outcome, the SARS-CoV-2 viral assay negative conversion time, we firstly used Wilcoxon rank sum test to test the difference of SARS-CoV-2 viral assay negative conversion time between treatment group and control group, and gave the medians and IQR (interquartile range). Then, kaplan-Meier method was used to estimate the cumulative negative conversion rate and draw the survival curve, and the negative conversion rate was compared among groups through log-rank test, the median time and 95% CIs (confidence interval) were given, and the hazard ratios was estimated by Cox regression model. Hazard ratios greater than 1 indicated that the negative conversion rate of treatment group was higher than that of control group.

For secondary outcomes, the change of 7-point scale was tested by Wilcoxon rank sum test. The rate of critical aggravation was tested by Pearson chi-square test. Both blood routine and blood biochemistry outcomes were numerical data, which was tested by t-test for normal distribution data and Wilcoxon rank sum test for non-normal distribution data. Fever recovery time was tested by Wilcoxon rank sum test. For symptom improvements, which meant the disappearance rate of cough, chest tightness, headache, fatigue, and pharynx, we used the Pearson chi-square test. For the chest CT outcome, the score of area and density for both ground glass and consolidation imaging manifestation, and overall evaluation of chest CT examination, were tested by Wilcoxon rank sum test.

Safety analyses were based on the patients’ actual exposure to treatment. In this study, statistical packages base, Stats, Rcompanion and survival in R version-3.6.2 and SPSS 20.0 were used for statistical analysis and verification.

### Patient and public involvement

No patients were involved in the research design and implementation plan. There is no plan to disseminate the result of this trial to study participants.

## RESULT

### Patients

Of the 285 patients were assessed from February 27 to March 27, 81 did not meet the eligibility criteria. Remaining 204 patients were randomised with a ratio 1:1 to treatment group and control group. 195 patients were included in the FAS (Full Analysis Set) (99 in treatment group and 96 in control group) (Figure 1).The mean age was 54 years old, and 36.4% was male. Table 1 showed baseline demographic, clinical characteristics of patients in treatment group and control group.

**Table 1.** Baseline demographic and clinical characteristics of patients in FAS.

| Characteristics | Treatment group (n=99) | Control Group (n=96) | Total (n=195) |
| --- | --- | --- | --- |
| Median (IQR) age, years | 56.00 (48.50-62.00) | 56.00 (48.50-62.00) | 56.00 (48.50- 62.00) |
| **SEX** |  |  |  |
| Male | 36 (36.4%) | 37 (38.5%) | 73 (37.4%) |
| **Disease severity** |  |  |  |
| Mild and Moderate | 74 (74.7%) | 89(92.7%) | 163 (83.6%) |
| Severe | 25 (25.3%) | 7 (7.3%) | 32 (16.4%) |
| **Coexisting condition** |  |  |  |
| Hypertension | 63 (63.6%) | 54 (56.2%) | 117 (60.0%) |
| Hyperlipemia | 6 (6.1%) | 10 (10.4%) | 16 (8.2%) |
| Coronary heart disease | 7 (7.1%) | 3 (3.1%) | 10 (5.1%) |
| Thyroid disorder | 1 (1.0%) | 1 (1.0%) | 2 (1.0%) |
| Cerebral lacunar infarction | 1 (1.0%) | 3 (3.1%) | 4 (2.1%) |
| Diabetes | 18 (18.2%) | 9 (9.4%) | 27 (13.8%) |
| Rheumatoid disease | 2 (2.0%) | 1 (1.0%) | 3 (1.5%) |
| Chronic bronchitis | 1 (1.0%) | 1 (1.0%) | 2 (1.0%) |
| **Symptoms** |  |  |  |
| Fever | 85 (85.9%) | 90 (93.8%) | 175 (89.7%) |
| Cough | 86 (86.9%) | 73 (76.0%) | 159 (81.5%) |
| Fatigue | 84 (84.8%) | 87 (90.6%) | 171 (87.7%) |
| Headache | 14 (14.1%) | 12 (12.5%) | 26 (13.3%) |
| Chest discomfort | 54 (54.5%) | 59 (61.5%) | 113 (57.9%) |
| Sore throat | 87 (87.9%) | 87 (90.6%) | 174 (89.2%) |
| **Laboratory parameters—Median(IQR)/mean (SD)** |  |  |  |
| White blood cell count, $\times 10^9/L$ | 5.84 (4.90- 7.29) | 5.68 (4.78- 6.75) | 5.77 (4.82- 6.89) |
| Red blood cell count, $\times 10^{12}/L$ | 4.13 (3.84- 4.44) | 4.04 (3.67- 4.35) | 4.06 (3.76- 4.39) |
| Platelet count, $\times 10^9/L$ | 229.00 (172.00- 267.50) | 212.00 (184.75- 261.75) | 222.00 (179.50- 265.50) |
| Hemoglobin, g/L | 125.00 (118.00- 135.50) | 122.50 (111.00- 133.50) | 124.00 (116.00- 135.00) |
| Neutrophil count, $\times 10^9/L$ | 3.66 (2.69- 4.43) | 3.28 (2.61- 4.19) | 3.42 (2.67- 4.35) |
| Lymphocyte count, $\times 10^9/L$ | 1.73 (1.48- 2.02) | 1.65 (1.38- 2.07) | 1.68 (1.42- 2.05) |
| Monocyte count, $\times 10^9/L$ | 0.41 (0.35- 0.49) | 0.38 (0.32- 0.46) | 0.40 (0.34- 0.48) |
| Neutrophil percentage, % | 59.30 (54.35- 65.55) | 58.85 (50.53- 64.10) | 59.10 (52.60- 65.20) |
| Lymphocyte percentage, % | 30.17 (8.41) | 30.27 (9.54) | 30.22 (8.96) |
| Monocyte percentage, % | 7.20 (6.10- 7.80) | 6.80 (5.88- 7.85) | 7.00 (6.00- 7.80) |
| Total bilirubin, $\mu\text{mol/L}$ | 12.20 (10.00- 15.55) | 11.90 (9.80- 14.88) | 12.10 (9.85- 15.25) |
| Alanine transaminase, U/L | 25.00 (19.00- 37.50) | 27.50 (19.00- 35.00) | 26.00 (19.00- 35.00) |
| Glutamic oxalacetic transaminase, U/L | 25.00 (22.00- 31.00) | 29.00 (23.00- 34.00) | 26.00 (22.00- 33.00) |
| Albumin, g/L | 39.40 (37.25- 40.85) | 39.10 (37.20- 40.50) | 39.20 (37.20- 40.70) |
| Urea, mmol/L | 4.50 (3.65- 5.23) | 4.57 (3.60- 5.50) | 4.54 (3.60- 5.37) |
| Creatinine, umol/L | 62.40 (53.50- 73.90) | 64.60 (56.50- 73.10) | 63.00 (54.65- 73.90) |
| Blood glucose, mmol/L | 5.20 (4.80- 6.05) | 5.10 (4.80- 6.12) | 5.20 (4.80- 6.10) |
| Creatine kinase, U/L | 63.00 (45.25- 85.75) | 65.50 (50.00- 95.00) | 63.50 (48.00- 87.00) |
| Creatine kinase isoenzyme, U/L | 10.00 (8.00- 12.00) | 11.00 (8.00- 12.25) | 10.00 (8.00- 12.00) |
| Blood potassium, mmol/L | 4.20 (4.00- 4.50) | 4.30 (4.00- 4.50) | 4.30 (4.00- 4.50) |
| Blood sodium, mmol/L | 140.00 (139.00- 141.00) | 140.00 (138.00- 142.00) | 140.00 (138.00- 142.00) |
| Blood chlorine, mmol/L | 106.00 (104.00- 108.00) | 106.00 (104.00- 107.00) | 106.00 (104.00- 107.00) |
| C-reactive protein, mg/L | 1.00 (0.50- 2.90) | 1.00 (0.60- 1.70) | 1.00 (0.50- 2.38) |
| Hypersensitive C-reactive protein, mg/L | 1.10 (0.50- 2.00) | 1.10 (0.50- 2.80) | 1.10 (0.50- 2.35) |
| Amyloid protein, mg/L | 3.70 (3.70- 6.45) | 3.70 (3.70- 5.75) | 3.70 (3.70- 6.15) |
| Myoglobin, ng/mL | 30.60 (24.70- 42.40) | 32.40 (24.95- 41.85) | 31.00 (24.80- 41.92) |
| Hypersensitive troponin, pg/mL | 1.50 (0.70- 3.40) | 1.70 (0.70- 3.20) | 1.70 (0.70- 3.40) |
| Blood sedimentation, mm/h | 15.00 (7.00- 23.00) | 16.00 (11.00- 25.00) | 16.00 (9.00- 25.00) |
| Ferritin, ng/mL | 196.43 (128.24- 309.99) | 195.99 (114.96- 300.11) | 195.99 (119.33- 309.75) |
| D-dimer, ug/mL | 0.38 (0.23- 0.65) | 0.44 (0.24- 0.84) | 0.40 (0.24- 0.76) |
| Interleukin 6, pg/mL | 7.02 (5.46- 8.59) | 6.78 (5.73- 8.43) | 6.94 (5.54- 8.54) |
| Procalcitonin , ng/ml | 0.05 (0.05- 0.05) | 0.05 (0.05- 0.05) | 0.05 (0.05- 0.05) |
| **Chest CT parameters—Median(IQR)** |  |  |  |
| score of ground glass area | 16.50 (10.25-21.75) | 17.00 (8.00-27.50) | 16.75 (9.00-24.12) |
| Score of consolidation area | 0.00 (0.00- 0.00) | 0.00 (0.00-0.00) | 0.00 (0.00-0.00) |

**Figure 1.**
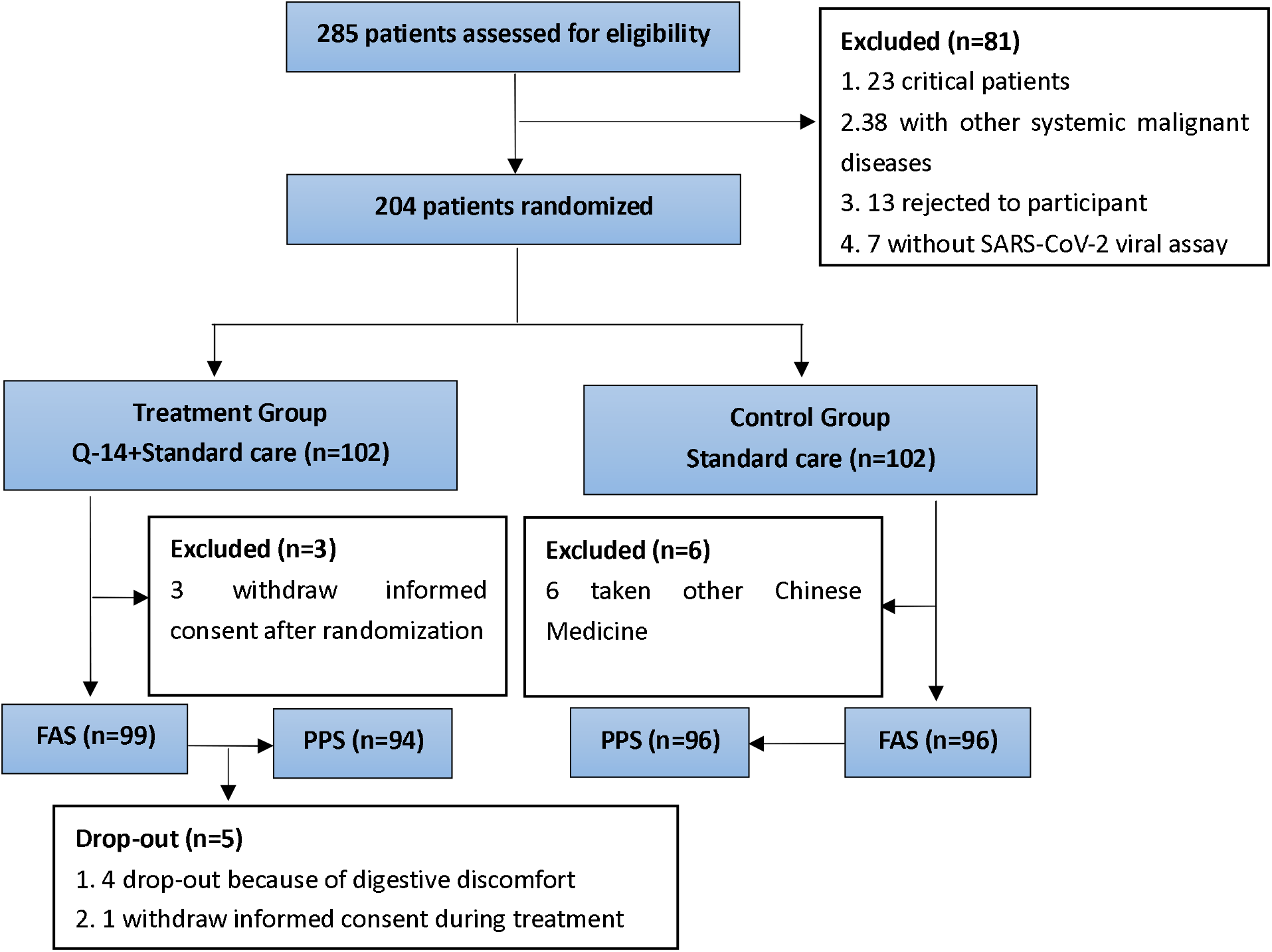
Research flow chart.

### Primary outcome

Among 195 patients in FAS, 149 (76.4%) patients (71 treatment group, 78 control group) had converted to negative before the cut-off day of analysis, and the remaining 46 patients (28 treatment group, 18 control group) did not arrive a conversion-to-negative result.

In the conversion time of SARS-CoV-2 viral assay, there was no significant difference between treatment group and control group both in FAS and PPS (Per-protocol Set) (FAS: n=149. Median (IQR): 10.00 (9.00-11.00) vs. 10.00 (9.00-11.00); Mean rank: 67.92 vs. 81.44; P=0.051. PPS: n=148. Median (IQR): 10.00 (9.00, 11.00) vs. 10.00 (9.00, 11.00); Mean rank: 67.86 vs. 80.46; P=0.068). The average time of conversion-to-negative showed some advantage in treatment group (Mean (SD): FAS: 10.01 (1.74) vs. 10.44(1.49) days. PPS: 10.04 (1.73) vs. 10.44(1.49) days).The median time to conversion between treatment group and control group showed no significant difference as well (FAS: 10.00 (9.00-10.00) vs. 10.00 (10.00-11.00) days, HR: 1.165, 95%CI: 0.84-1.61, P=0.27. PPS: 10.00 (9.00-11.00) vs. 10.00 (10.00-11.00) days, HR = 1.15, 95%CI: 0.83-1.59, P=0.31 by log rank test. Figure 2, Figure 3).

**Figure 2.**
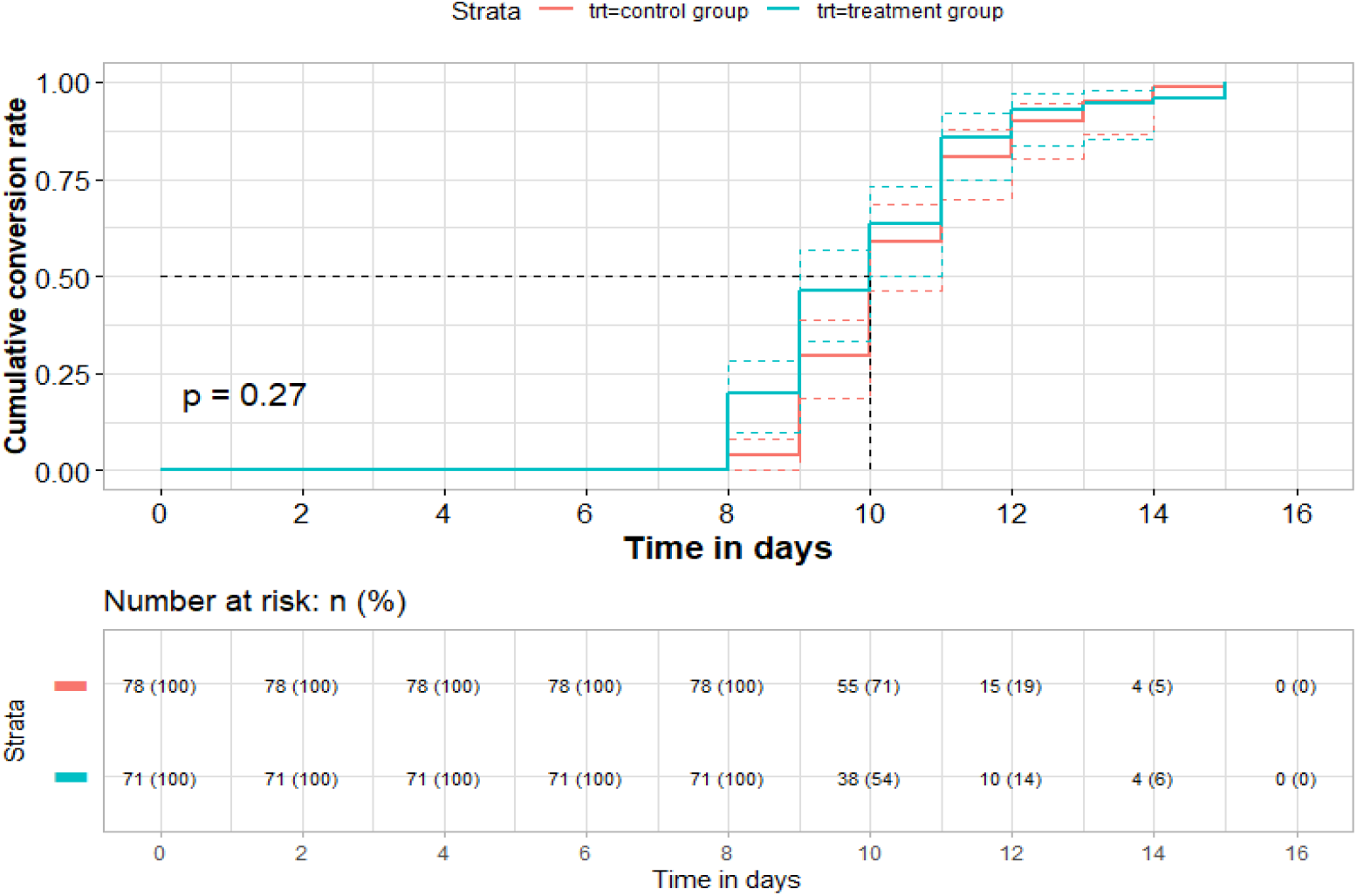
Time to conversion analysis of SARS-CoV-2 viral assay and conversion ending (n=149, FAS)

**Figure 3.**
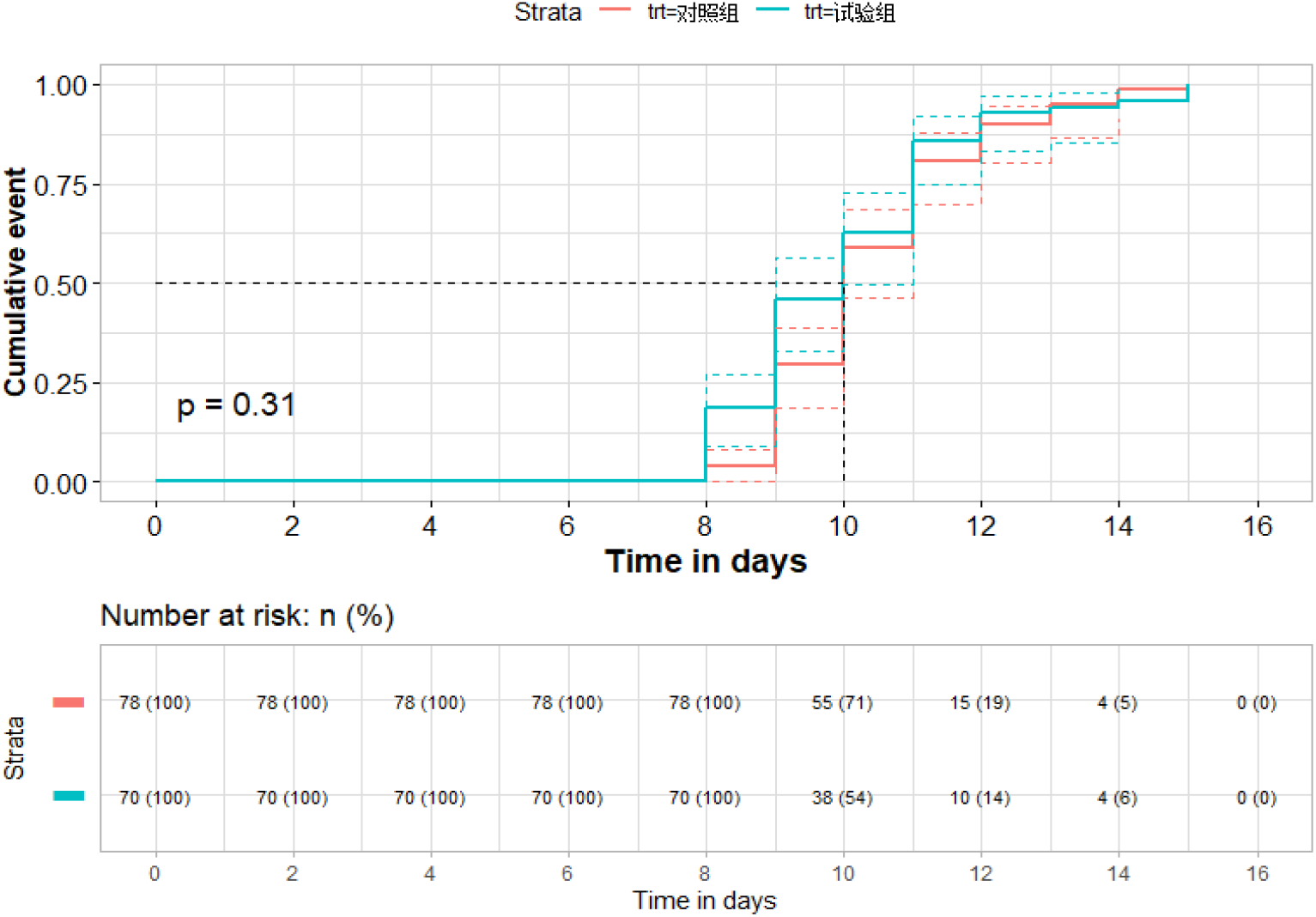
Time to conversion analysis of SARS-CoV-2 viral assay and conversion ending (n=148, PPS)

We found among the 116 participants with cardiovascular disease who reached conversion-to-negative, the conversion time of SARS-CoV-2 viral assay is significantly shorter in treatment group as compared with control group.(Median (IQR): 10.00 (9.00-11.00) vs. 10.00 (9.00-11.00); Mean rank: 51.63 vs. 65.61; P=0.022.). The average time of conversion-to-negative showed some advantage in treatment group (Mean (SD): FAS: 9.85 (1.42) vs. 10.54 (1.56) days). The median time to conversion between treatment group and control group showed significantly difference (10.00 (9.00-10.00) vs. 10.00 (10.00-11.00) days, HR: 1.484, 95%CI: 1.02-2.15, P=0.023 by log rank test. Figure 4).

**Figure 4.**
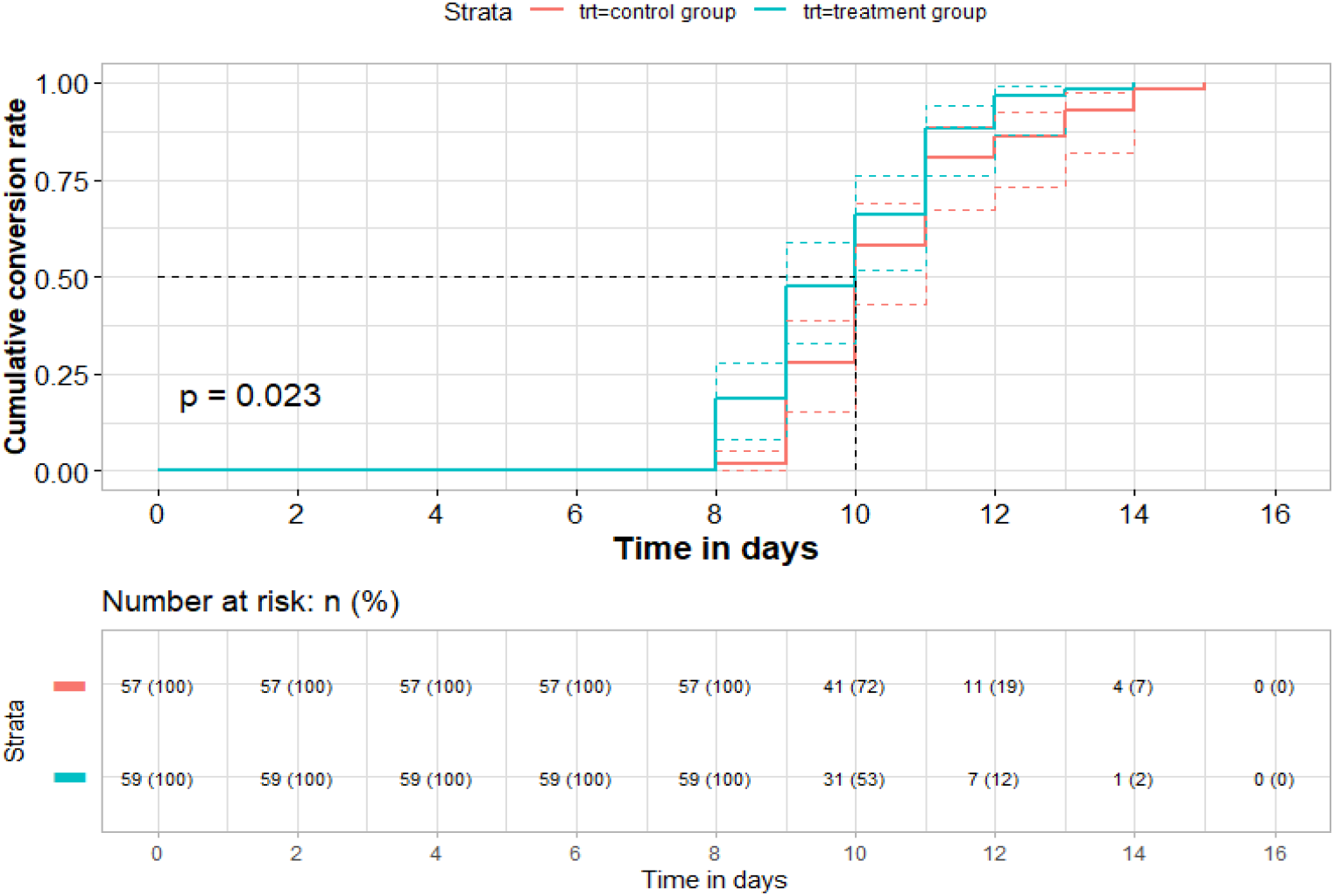
Time to conversion analysis of SARS-CoV-2 viral assay and conversion ending, in participants with cardiovascular disease who reached negative conversion (n=116)

### Secondary outcome

The change of the 7-point scale was similar in the two groups (Median (ICQ): FAS: -2.00(-3.00-0.00) vs. -2.00(-3.00-0.00), P=0.109. PPS: -2.00(-3.00, 0.00) vs. -2.00(-3.00, 0.00), P=0.232).

There was no patient turned critical category during the observation.

There was no significant difference in blood routine test and blood biochemical test.

Fever recovery time was shorter in treatment group as compared in control group (Median (ICQ): FAS: 2.00 (2.00-3.00) vs. 3.00 (1.00-4.00) days, P=0.029. PPS: 2.00 (2.00-3.00) vs. 3.00 (1.00-4.00), P=0.049).

The symptom disappearance rate of cough (FAS: 90.7% vs. 79.5%, P=0.045. PPS: 91.7% vs. 79.5%, P=0.028), fatigue (FAS: 88.1% vs. 75.9%, P=0.038. PPS: 87.8% vs. 75.9%, P=0.045), chest discomfort (FAS: 87.0% vs. 67.8%, P=0.015. PPS: 88.5% vs. 67.8%, P=0.009) was significant higher in treatment group. The symptom disappearance rate of headache (FAS and PPS: 85.7% vs. 83.3%, P> 0.999) and sore throat (FAS: 82.8% vs. 74.7%, P=0.195. PPS: 81.7% vs. 74.7%, P=0.272) was similar in the two groups.

In chest computed tomography (Chest CT) examinations, the score of ground glass area was significantly lower in treatment group than control group after treatment(Median (ICQ):FAS: 8.50 (5.00-15.25) vs. 14.00 (7.00-21.75), P=0.031. PPS: 9.00 (4.62-16.00) vs. 14.00 (7.00-21.75), P=0.042)with a larger decrease (Median (ICQ):FAS: -5.75 (-11.25- -2.00) vs.-0.75 (-2.12-2.12), P< 0.001. PPS: -6.50 (-11.75--2.00) vs.-0.75 (-2.12-2.12), P< 0.001). The density of ground glass area showed a significant decrease in treatment group (FAS: decreased 36 (80.0%), no-change 9(20.0%), increased 0 (0%) vs. decreased 17 (51.5%), no-change 15(45.5%), increased 1(3.0%), P=0.007.PPS: decreased 33 (78.6%), no-change 9(21.4%), increased 0 (0%) vs. decreased 17 (51.5%), no-change 15(45.5%), increased 1(3.0%), P=0.013.)Overall evaluation of chest CT examination after treatment compared with baseline showed more patients improved in treatment group (FAS: improved69 (85.2%), no-change 12(14.8%), aggravated 0 (0%) vs. improved55 (67.9%), no-change 23(28.4%), aggravated 1(3.7%), P=0.008.PPS: improved65 (85.5%), no-change 11(14.5%), aggravated 0 (0%) vs. improved 55 (67.9%), no-change 23(28.4%), aggravated 1(3.7%), P=0.008.)There was no significant difference in score of area and density of consolidation manifestation.

### Safety

Among 204 patients, a total of 202 cases were analyzed as the safety population (100 treatment group, 102 control groups), as there were two patients who were assigned to treatment group withdraw their informed consent on the day of randomization and having no baseline information. The comparison of recorded adverse events showed there were 14 patients (16 adverse events) in treatment group and 15 patients (23 adverse events)) in control group (Table 2). There were no serious adverse events recorded. Diarrhea (n=15) was the most common adverse event in this trial. For patients who happened digestive discomfort, the Q-14 dosage in treatment group adjusted to half dosage daily until the symptom improved. Other adverse events were transient with duration of one to two days.

**Table 2.** Summary of adverse events in safety population*.

| Adverse events | Treatment group (n=100) | Control Group (n=102) | Total (n=202) |
| --- | --- | --- | --- |
| **All adverse even** | **16** | **23** | **35** |
| Diarrhea | 8 (8%) | 7 (6.9%) | 15 (7.4%) |
| Abdominal discomfort | 2 (2%) | 7 (2%) | 5 (2.5%) |
| Decreased appetite | 3 (3%) | 0 (0) | 3 (1.5%) |
| Anxiety | 0 (0) | 2 (2.0%) | 2 (1.0%) |
| Oral ulcer | 1 (1%) | 1 (1.0%) | 2 (1.0%) |
| short breath | 0 (0) | 2 (2.0%) | 2 (1.0%) |
| Constipation | 0 (0) | 1 (1.0%) | 1 (0.5%) |
| Vomiting | 1 (1%) | 0 (0) | 1 (0.5%) |
| Itchy skin | 0 (0) | 1 (1.0%) | 1 (0.5%) |
| Lower extremity edema | 0 (0) | 1 (1.0%) | 1 (0.5%) |
| Dry eye | 1 (1%) | 0 (0) | 1 (0.5%) |
| Limb pain | 0 (0) | 1 (1.0%) | 1 (0.5%) |

\*Values stand for numbers (percentages of patient happened adverse events)

## DISSCUSION

This clinical trial (conducted during the outbreak of COVID-19 in Wuhan, China) is the first registered randomised controlled trial evaluating administration of Q-14 in hospitalized adult patients with laboratory confirmed COVID-19 in Wuhan, China. These finds provide evidence to support a notable improvement of typical uncomfortable symptoms caused by COVID-19, such as fever, cough, fatigue and chest discomfort. Additional, a significant improvement of Chest CT by a 14-day course of Q-14 administration to the standard care in hospitalized adult patients with COVID-19.

At present, there is no definite antiviral drug to accelerate the probability of negativeconversion for SARS-CoV-2 (Parvathaneni and Gupta, 2020; Yadav et al, 2020), the development of a vaccine (Torreele, 2020; Wang et al., 2020) appears to be the last straw for prevention and control of COVID-19. The development of new antiviral requires a considerable length of time and effort for drug design and validation. Clearly, not only will this take time, but everything is unknown. Administration of Q-14 did not resultin a significantly higher probability of negativeconversion than standard of care alone in patientsadmitted to hospital with COVID-19, but it is beneficial for the improvement of typical symptoms in COVID-19 patients. Improvement in clinical symptoms is important to reduce the level of discomfort in hospitalized adult patients with COVID-19, at the same time; it is beneficial to relieve depressive or anxiety caused by typical symptoms of COVID-19 patients or medical workers (Elhadi et al., 2020; Wang et al., 2020), such as fever, cough, fatigue and chest discomfort. In our study, the safety of Q-14 has been fully investigated. No serious adverse events were reported except for diarrhea (8 cases of treatment group) or other adverse reactions in some patients, which resolve themselves within a few days. The overall clinical safety of a 14-day course of Q-14 administration is documented.

On the other hand, we performed a subgroup analysis, an increase the probability of negative conversion of SARS-CoV-2 viral assay conferred by the addition of a 14-day course of Q-14 administration to the standard care in hospitalized adult patients with COVID-19 and cardiovascular diseases. Although the results were statistically significant, they were not particularly significant in terms of time (days). This is not within our expectation, which is an interesting finding. Although we cannot rule out the possibility of a chance finding, we think it may be related to the following factor. The majority of COVID-19 patients with cardiovascular diseases are hypertensive; Q-14 may have a certain targeted improvement effect on the pathological status of hypertension addition of standard care. Molecular docking showed that baicalein and quercetin were the top two compounds of Q14, baicalein had strong affinity for SARS-CoV-2, and quercetin had strong affinity forACE2 3CL that indicated that baicalein and quercetin might play an important role in the treatment of SARS-CoV-2 (Tao et al., 2020) .Future clinical studies with a larger sample are needed to confirm this effect of Q-14. COVID-19 patients with pre-existing cardiovascular diseases experience disproportionately worse outcomes in mortality (Peng et al., 2020; Golemi et al., 2020), the findings of this clinical study may provide an effective drug for the prevention and treatment of COVID-19 patients with hypertension. Of course, more rigorous clinical and basic research is needed to validate the speculation.

Chest CT is used for diagnosis of COVID-19, as an important complement to the RT-PCR (reverse-transcription polymerase chain reaction) tests (Ai et al., 2020; Wong et al., 2020). In the early stages of the COVID-19 epidemic, chest CT may be of greater diagnostic value than RT-PCR tests. At the same time, it will be a very important observational indicator to evaluate the curative effect for therapeutic schedule. Not only that, scientists have proposed a non-invasive and quantitative prognostic tool for predicting poor outcome in patients with COVID-19 based on CT imaging (Wu et al.,2020). Our study found that after 14 days of treatment, Q-14 with standard care can significantly improve abnormal chest CT changes of COVID-19 patients, decreased the score of ground glass area (characteristic chest CT change of COVID-19) significantly. This result suggests that Q-14 has an important therapeutic role in alleviating the condition of COVID-19 patients.

More and more clinical evidence shows the great role of TCM in the prevention and control of COVID-19 (Hu et al., 2020; Xiao et al., 2020; Xiong et al., 2020). From these studies, we have not seen that Traditional Chinese medicine has a direct antiviral effect in clinical practice, but its ability to quickly relieve clinical symptoms may be closely related to the enhancement of the immune ability of the body.

## LIMITATIONS OF STUDY

Some limitations existed in the present study. First, we did not set up a placebo as a Q-14 control. In the context of the COVID-19 early outbreak in Wuhan, China, many clinical trials related COVID-19 has yet to be better prepared because of Wuhan quarantine, especially for Q-14 placebo. As we did not adopt stratify randomisation according to disease severity, the cases of severe patient showed significant difference between the two groups. This may have greatly influenced the conclusions of this trial, placebo effect of Q-14 cannot be ruled out. Secondly, the open label rather than double blind design, which may be resulted in possibility of biased assessments. Thirdly, we began to determine SARS-CoV-2 RNA conversion from the 7th day of the intervention, which may have overlooked some patients who have negative conversion of SARS-CoV-2 within 7 days. Finally, the specimens collected in our trial for SARS-CoV-2 RNA determination were mostly from the upper respiratory tract rather than bronchoalveolar lavage fluid, which could result in false negative results.

## CONCLUSION AND POLICY IMPLICATIONS

The introduction of TCM into the world is a demand of The Times, which is the worldwide urgent demand for TCM. This trial provides evidence to support a notable improvement of typical uncomfortable symptoms caused by COVID-19, such as fever, cough, fatigue and chest discomfort. Additional, a significant improvement of Chest CT by the addition of a 14-day course of Q-14 administration to the standard care in hospitalized adult patients with COVID-19. In the current situation, Q-14 may provide safe and effective option for controlling the spread of COVID-19, or a solution with Chinese characteristics. China’s great success in prevention and control of COVID-19 has demonstrated the feasibility of Chinese Approach.

## Supporting information

composition of Q-14

EMERGENCY PRODUCT REGISTRATION CERTIFICATE

## Data Availability

Research data was recorded using Medroad Cloud Electronic Data Capture System (Jiangsu Famous Medicine Science and Technology Ltd.) http://cloud.medroad.cn/

## FUNDING

The study was funded by National Key Research and Development Plan for the Emergency Management of Novel Coronavirus Pneumonia (No. 2020YFC0841500)

## AUTHOR’S CONTRIBUTION

JL,WY contributed equally to this paper and share joint first authorship. JL contributed to the writing on this paper and in charge of the trial management during this study. WY contributed to the data cleaning and statistical analysis. YL contributed to the result analysis and modified the study report. CL contributed to the data collection organization on-site in Wuhan Jinyint Hospital and carried out the first level quality control. LR contributed to the organization in Wuhan Jinyintan Hospital and communicated with the patients. CZ designed the protocol and finished the trial registration. RH contributed to the data cleaning and assisted the trial management. XS led the data collection and data recording. QM, WL, HL contributed as clinical expert on-site in Wuhan Jinyintan Hospital from protocol designing to implementation. HS contributed to the trial drug coordinating on-site in Wuhan. YC,ZZ contributed to randomisation number application. LH, ZY, LZ, BW, GD, YX, BL, CX, YC, YB, JG, JY, JW, WQ, SC, BY, WW, JL, XX, MX, JJ, GW, XC contributed to theimplementation of intervention during this trial.HZ, JS, LL, DL contributed to collect data on-site in Wuhan. LG contributed to finalize the structure of this paper. LH organized the research group and contributed to the overall designing and management of this trialfor the corresponding authorship.

## ACKNOWLEDGEMENTS

We thank China National Medical Team of Traditional Chinese Medicine of China Academy of Chinese Medical Sciences (CNMTCM-CACMS Team), who went to Wuhan in the most critical moment for medical aid and with their hard working, we finished this trial. The member of CNMTCM-CACMS Team including Luqi Huang, Cheng Lv, Qing Miao, Wenliang Lv, Hao Li, Huaxin Shi, Lijie Hu, Zhixu Yang, Li Zhang, Bing Wang, Guoju Dong, Yongyue Xian, Bin Li, Zhenqi Zhou, Chunyan Xu, Yingying Chen, Yongjun Bian, Jing Guo, Jinliang Yang, Jian Wang, Wensheng Qi, Suping Chen, Yang Chen, Bei Yan, Wei Wang, Jing Li, Xiaolei Xie, Ming Xu, Jianxin Jiang, Gang Wang, Xiaodong Cong, Haoning Zhu, Jiaheng Shi, Luxing Leng, Dongxu Li. Xinghua Xiang and Yunfei Xing assisted the statistical work. Mingjiang Yao, Ping He, Xiao Liang, Ruiyan Cao, Li Li, Benliang Zou, Ning Wang, Shihuan Tang, Min Li, Xunlu Yin, Hao Wang, Chuanzhou Yang, Chunyu Gao, Liying Wang, Yongli Dong, Ming Chen, Lu Zhang, Xu Wei contributed to the data recording.Yang Zhao, Qiaoning Yang, Rui Li,Yingjue Jia,Yu Dong carried out the second level quality control. Hongjun Yang carried out the third level quality control and Yanping Wang contributed to the management of the trial. Xuedong Yang and Chunzhi Li contributed to the CT reports and images review. Meng Wu contributed to the CT review data verifying.

## DECLARATION OF COMPETING INTEREST

None.

## EXCLUSIVE LICENCE

The Corresponding Author has the right to grant on behalf of all authors and does grant on behalf of all authors, a worldwide licence to the Publishers and its licensees in perpetuity, in all forms, formats and media according to the requirement on exclusive licence of the publication.

## DATA AVAILABILITY STATEMENT

The data of this article could be shared within six months after the trial complete via the Medroad Cloud Electronic Data Capture System (http://cloud.medroad.cn/).

## Abbreviations

CI: Confidence interval
CNMTCM-CACMS: China National Medical Team of Traditional Chinese Medicine of China Academy of Chinese Medical Sciences
CONSORT: Consolidated Standards of Reporting Trials
COVID-19: Coronavirus Disease 2019
CT: Computed tomography
ECMO: Extracorporeal membrane oxygenation
FAS: Full Analysis Set
GCP: Good Clinical Practice
IQR: Interquartile range
NHC-NATCM-China guidelines: National clinical practice guideline for COVID-19 in China” developed by the National Health Commission and the National Administration of TCM of the People’s Republic of China
PPS: Per-protocol Set
RT-PCR: Reverse-transcription polymerase chain reaction
SCIO-China: State Council Information Office of the People’s Republic of China
TCM: Traditional Chinese medicine
WHO: World Health Organization

---

**Annex 1.**
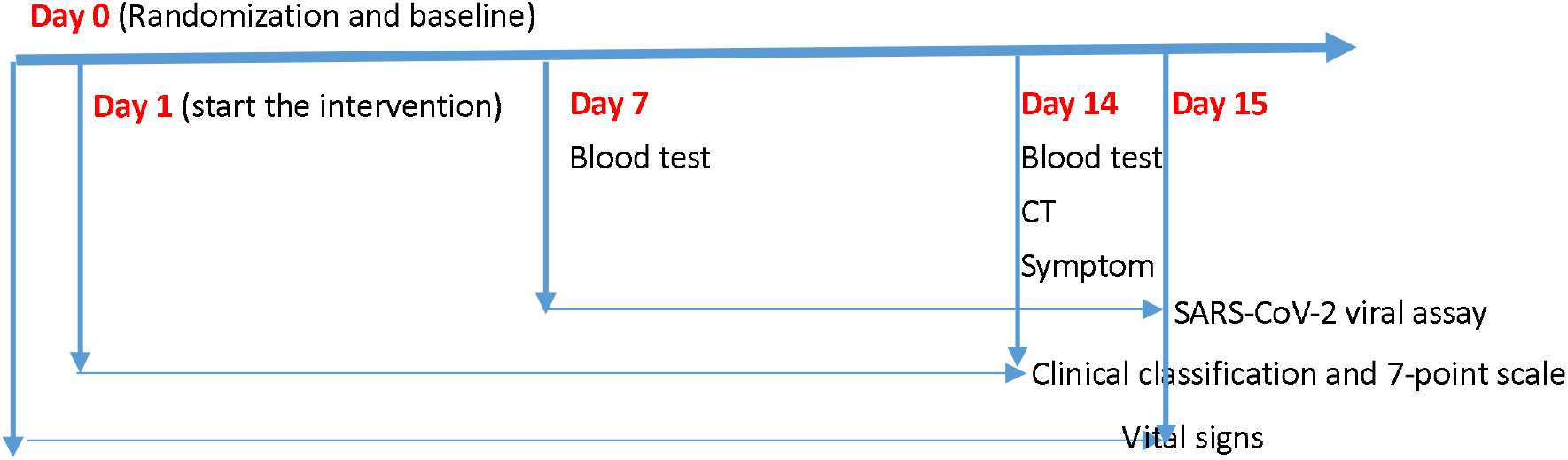
Research Period.

## Notes

### Competing Interest Statement

The authors have declared no competing interest.

### Clinical Trial

ChiCTR2000030288

### Author Declarations

The study protocol had been approved by the Ethics Review Committee of China Academy of Chinese Medical Sciences.

