## Supplementary material for "Chinese medicine (Q-14) in the Treatment of Patients with Coronavirus Disease 2019 (COVID-19): A Single-center, Open label, Randomised Controlled Trial": composition of Q-14

***Q-14* is consisted of thirteen herbal and one mineral medicines**:

Mahuang (Herba Ephedrae), 6 g;

Kuxingren (Armeniacae Semen Amarum), 9 g;

Shigao (Gypsum Fibrosum), 15 g;

Gancao (Glycyrrhizae Radix et Rhizoma), 3 g;

Huoxiang (Pogostemonis Herba), 10 g;

Houpu (Magnoliae Officinalis Cortex), 10 g;

Cangzhu (Atractylodis Rhizoma), 15 g;

Caoguo (Tsaoko Fructus), 10 g;

Fabanxia (Pinelliae Rhizoma Praeparatum), 9 g;

Fuling (Poria), 15 g;

Shengdahuang (Rhei Radix et Rhizoma), 5 g;

Shenghuangqi (Astragali radix), 10 g;

Tinglizi (Descurainiae Semen Lepidii Semen), 10 g;

Chishao (Paeoniae Radix Rubra), 10 g.
