## Supplementary material for "Chinese medicine (Q-14) in the Treatment of Patients with Coronavirus Disease 2019 (COVID-19): A Single-center, Open label, Randomised Controlled Trial": EMERGENCY PRODUCT REGISTRATION CERTIFICATE

Issued by Ministry of Health & Prevention of United Arab Emirates

Certificate #: **DR/C/2020/10-C1**

Issued on: 01/09/2020

**Herbal Medicines**

*It is certified that the following product was approved for emergency temporary registration in the Ministry of Health and Prevention.*

| **Registration No.** | DR/C/2020/10 | **First Reg. Date:** | 01/09/2020 |
| --- | --- | --- | --- |
| **Expiry Date:** | 31/08/2021 | **Product Name** | Huashi Baidu |
| **Pharmaceutical Form** | Granules |  |  |
| **Dispensing Mode** | | **Prescription Only Medicine (POM)** | |
| **Active Ingredient(s)** | | **Manufacturer** | |
| Ephedra Patchouli Raw Gypsum Fried Bitter Almonds Pinellia Magnolia Bran Fried Atractylodes Fried Grass Nuts Poria Astragalus Red Peony Draba nemorosa L, Rhubarb, Licorice | | Guangdong Yifang Pharmaceutical Co.,ltd [Guangdong;CHINA] | |
|  |  | **Marketing Authorization Holder** | |
|  |  | Guangdong Yifang Pharmaceutical Co.,ltd [Guangdong;CHINA] | |
|  |  | **Local Agent** | |
|  |  | G42 MEDICATIONS TRADING LLC. [ABU DHABI; UAE] | |
